# The Cartilage Thickness Score (CTh-Score) detects a structural effect of 2-year weight loss: data from the Osteoarthritis Initiative

**DOI:** 10.64898/2026.04.01.26349854

**Authors:** Paul Margain, Julien Favre, Francis Berenbaum, Patrick Omoumi

## Abstract

**Objective:** To determine whether clinically significant weight loss (≥5% of body weight) is associated with slower 2-year knee cartilage structural deterioration, using the Cartilage Thickness Score (CTh-Score).

**Method:** A retrospective propensity-score-matched cohort study was conducted using Osteoarthritis Initiative study data. Weight loss was defined as ≥5%; stable weight as <5% loss and <5% gain. Cases were matched 1:2 to controls using age, sex, body mass index, visit, Kellgren–Lawrence grade, KOOS Pain, and physical activity. The primary outcome was change in CTh-Score, a 0-100 cartilage structural assessment score derived from the spatial distribution of cartilage thickness, with higher scores indicating greater severity. Secondary outcomes were KOOS Pain and conventional structural metrics: minimum medial joint space width and mean cartilage thickness in four femoral/tibial regions. Linear regression with matched-set cluster-robust standard errors estimated crude and adjusted differences with 95% confidence intervals (CIs).

**Results:** A total of 643 weight-loss participants were matched to 1,286 stable-weight controls; all absolute standardized mean differences were <0.10. Weight loss participants experienced less CTh-Score progression over two years than stable-weight controls (mean change 2.71 (SD 6.85) versus 4.29 (8.00), respectively). The adjusted between-group difference (weight loss minus stable weight) was −1.59 (95% CI −2.29 to −0.89; p<0.001; Cohen’s d=−0.21). There was no evidence that the association differed by radiographic osteoarthritis status (p=0.261). No statistically significant adjusted differences were found for minimum joint space width or regional mean cartilage thickness. The adjusted KOOS Pain difference was 1.14 points (95% CI −0.08 to 2.35; p=0.070).

**Conclusions:** Clinically significant weight loss was associated with slower knee cartilage structural progression over 2 years when assessed using the CTh-Score, despite no detectable differences in conventional structural measures. These findings suggest that weight management is associated with more favorable cartilage structural outcomes and highlight the CTh-Score as a sensitive endpoint for detecting structural change.

## Introduction

Osteoarthritis (OA) is a highly prevalent, chronic degenerative joint disorder characterized by progressive articular cartilage degradation, subchondral bone remodeling, and synovial inflammation, leading to pain and functional impairment [1–4]. The knee is the most commonly affected joint, making knee OA a leading cause of disability worldwide [5]. In the absence of approved disease-modifying osteoarthritis drugs capable of halting or reversing structural progression, current management strategies focus on alleviating symptoms and preserving joint function [1,6]. Among these strategies, weight loss constitutes a cornerstone recommendation for overweight or obese individuals with knee OA [7,8].

The benefits of weight loss are supported by evidence demonstrating that a reduction of at least 5% of body weight yields clinically meaningful improvements in pain and functional disability [9–12]. Furthermore, as obesity is a primary modifiable risk factor for incident knee OA, weight management also represents an effective preventive strategy [13]. The effects of weight loss are hypothesized to occur due to a dual mechanism: reducing biomechanical loading across the knee joint to alleviate cartilage stress, and lowering systemic low-grade inflammation driven by adipokines, both central to the pathophysiology of OA [4,13].

Although the symptomatic benefits of weight loss are well established, its impact on the structural integrity of articular cartilage remains uncertain. Previous investigations have yielded conflicting results; randomized trials with follow-up periods of up to 18 months reported no significant changes in traditional imaging measures of cartilage morphology on MRI or radiographs, such as radiographic minimal joint space width (JSW) or mean cartilage thickness within predefined regions, [10,14]. However, observational studies assessing changes in cartilage composition or expert lesion grading identified associations with weight loss over extended periods of four to eight years but did not assess short-term change [15,16]. This discrepancy raises a question: is the structural effect of weight loss only detectable over long term, or do traditional imaging metrics lack the sensitivity to detect changes within the shorter timeframes typical of clinical trials?

Recently, the Cartilage Thickness Score (CTh-Score), an automated MRI-based method, was developed to assess cartilage structural OA severity [17]. Utilizing deep learning to recognize spatial cartilage thickness patterns at high resolution, this method generates a continuous score from 0 to 100, with higher values indicating greater severity. It summarizes the entire tibiofemoral joint excluding the patella and is location-independent. Previous validations have demonstrated the CTh-Score’s high reproducibility (ICC > 0.98), generalizability beyond the OAI dataset and strong correlation with expert grading (r = 0.81, n=2671)[17]. Notably, the score has been shown to detect structural progression up to six years prior to radiographic changes [17]. By capturing the nuanced and spatially heterogeneous patterns of cartilage thickness distribution, the CTh-Score appears to represent a robust endpoint for assessing the structural effects of interventions such as weight loss. In the rest of the manuscript, structural progression refers to cartilage structural deterioration as assessed by changes in the CTh-Score.

Therefore, the objective of this study was to investigate the association between ≥5% weight loss and 2-year changes in knee cartilage structure, as assessed by the CTh-Score, in participants from the Osteoarthritis Initiative [18]. We hypothesized that individuals achieving ≥5% weight loss would demonstrate significantly less structural progression over two years than the control group.

## Methods

### Study Design and Population

This retrospective propensity-score-matched cohort study used the OAI, a multicenter prospective cohort of 4,796 adults aged 45–79 years with or at risk of symptomatic knee OA [18–20]. All participants gave written informed consent, and participating centers obtained institutional review board approval. Four exact two-year windows were considered (00–24, 24– 48, 48–72, and 72–96 months). In the rest of the manuscript, “baseline” refers to the first timepoint of the two-year window for each subject. For each window, clinical variables, Physical Activity Scale for the Elderly (PASE) and Knee injury and Osteoarthritis Outcome Score (KOOS) Pain data were obtained at its start and end; imaging variables were required at both visits[18,19,21].

### Clinical and Imaging Data Acquisition

For radiographic assessment, standardized fixed-flexion posteroanterior knee radiographs were acquired according to the OAI protocol. Kellgren-Lawrence (KL) grades and automated measurements of the minimum medial joint space width (JSW) were obtained from the central OAI database for this analysis [22–24]. Acquisitions were performed on 3.0T MRI using a standardized sagittal 3D dual-echo steady-state (DESS) sequence. Femorotibial bones and cartilage were automatically segmented from these scans using a previously validated deep learning algorithm [17] (Figure 1). These segmentations served to calculate mean cartilage thickness in four regions of interest (ROI); the medial femur, lateral femur, medial tibia, and lateral tibia. High-resolution cartilage thickness maps (CTh-Maps) along with their corresponding CTh-Score were extracted from a public repository [25].

**Figure 1.**
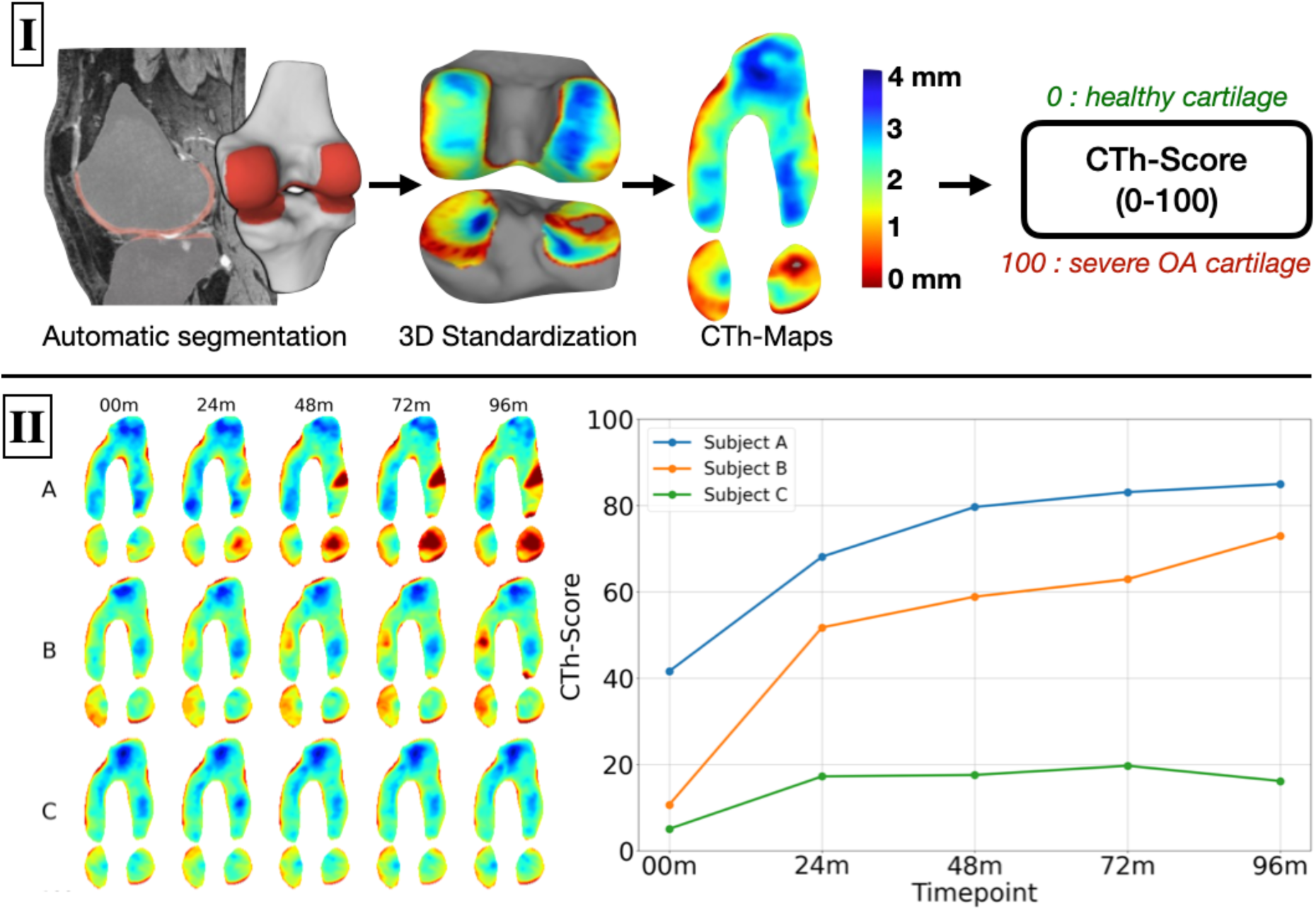
CTh-Score derivation and longitudinal representation. Panel I illustrates automatic femorotibial cartilage segmentation, three-dimensional standardization, generation of cartilage thickness maps (0–4 mm color scale), and conversion to the CTh-Score (0–100). The score summarizes location-independent spatial thickness patterns across the whole tibiofemoral joint, and higher scores indicate greater severity. Panel II shows longitudinal maps and scores for three illustrative participants (A–C) from 00 to 96 months. CTh, cartilage thickness.

### Participant Selection and Group Definition

Eligible participant–knee–window records had complete imaging and matching covariate data. Weight loss was defined as a decrease of ≥5% from the start of the interval, consistent with clinically meaningful thresholds used in prior knee OA trials [10,11]. Stable weight was defined as <5% loss and <5% gain; participants with ≥5% gain were excluded. For participants with more than one qualifying weight-loss interval, the earliest was selected. If both knees were eligible in that interval, the knee with the higher KL grade was selected. Participants who ever qualified as weight-loss cases were excluded from the control pool. Propensity scores were estimated by logistic regression using age, sex, body mass index (BMI), KL grade, PASE, KOOS Pain; continuous covariates were standardized. Cases were matched directly to two controls by nearest-neighbor matching without replacement, exactly for the OAI visit and radiographic OA status (KL<2 or KL≥2), using a caliper of 0.2 SD of the logit propensity score. Each participant appeared once in the final cohort.

### Statistical Analysis

The primary estimand was the adjusted mean difference (weight loss minus stable weight) in two-year CTh-Score change among matched participants. Balance was summarized using absolute standardized mean differences (SMDs), with <0.10 indicating good balance. Ordinary least-squares regression used cluster-robust standard errors at the matched-set level. The crude model included exposure group only. The adjusted model included baseline CTh-Score, age, sex, BMI, radiographic OA severity (KL), visit, KOOS Pain, and PASE. Adjusted marginal means and 95% confidence intervals (CIs) were calculated, and Cohen’s d was reported for effect size. Secondary outcomes, minimum medial JSW, mean cartilage thickness in the four regions, and KOOS Pain, were analyzed analogously with the corresponding baseline outcome; a positive KOOS Pain change indicates less pain. Effect modification by radiographic OA status was assessed secondarily by adding a weight-loss-by-OA interaction.

## Results

### Participant Characteristics

Of 4,796 OAI participants, 2,902 had at least one eligible two-year interval with complete data. The candidate pool included 690 participants with ≥5% weight loss and 2,126 with stable weight. Matching retained 643 weight-loss participants and 1,286 controls (Figure 2).

**Figure 2.**
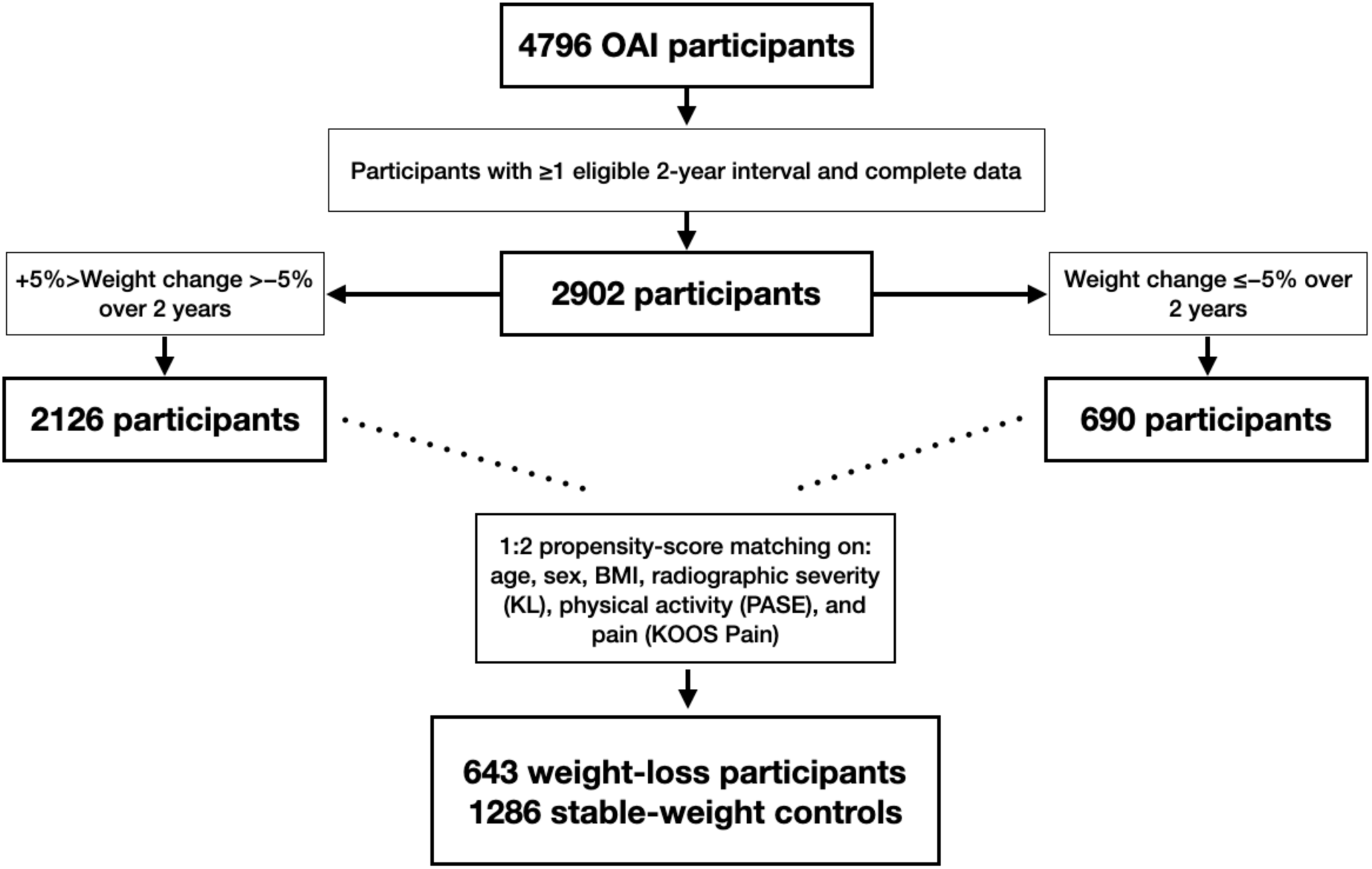
Participant selection and propensity-score matching. Four exact two-year windows were considered (00–24, 24–48, 48–72, and 72–96 months). For each participant, the earliest qualifying weight-loss interval was selected, and the index knee was the one with the highest KL grade. Stable weight was defined as <5% loss and <5% gain, and participants who ever met the weight-loss definition were excluded from the control pool. 86 participants were excluded because of weight gains ≥5% at each 2-year interval. Cases were matched directly 1:2 without replacement, exactly within window and radiographic OA severity, using start age, sex, BMI, KL grade, PASE, and KOOS Pain, with a 0.2-SD caliper on the logit propensity score. BMI, body mass index; KL, Kellgren–Lawrence; KOOS, Knee injury and Osteoarthritis Outcome Score; OA, osteoarthritis; OAI, Osteoarthritis Initiative; PASE, Physical Activity Scale for the Elderly; WL, weight loss.

Baseline characteristics were similar: mean CTh-Score was 35.9 (SD 24.4) versus 36.2 (24.0), and all absolute SMDs were <0.10. The weight-loss and stable-weight groups included 434 (67.5%) and 868 (67.5%) participants with radiographic OA, respectively. Mean age was 63.4 versus 63.3 years, 61.0% versus 59.8% were female, and mean BMI was 29.8 kg/m² in both groups (Table 1).

**Table 1.**
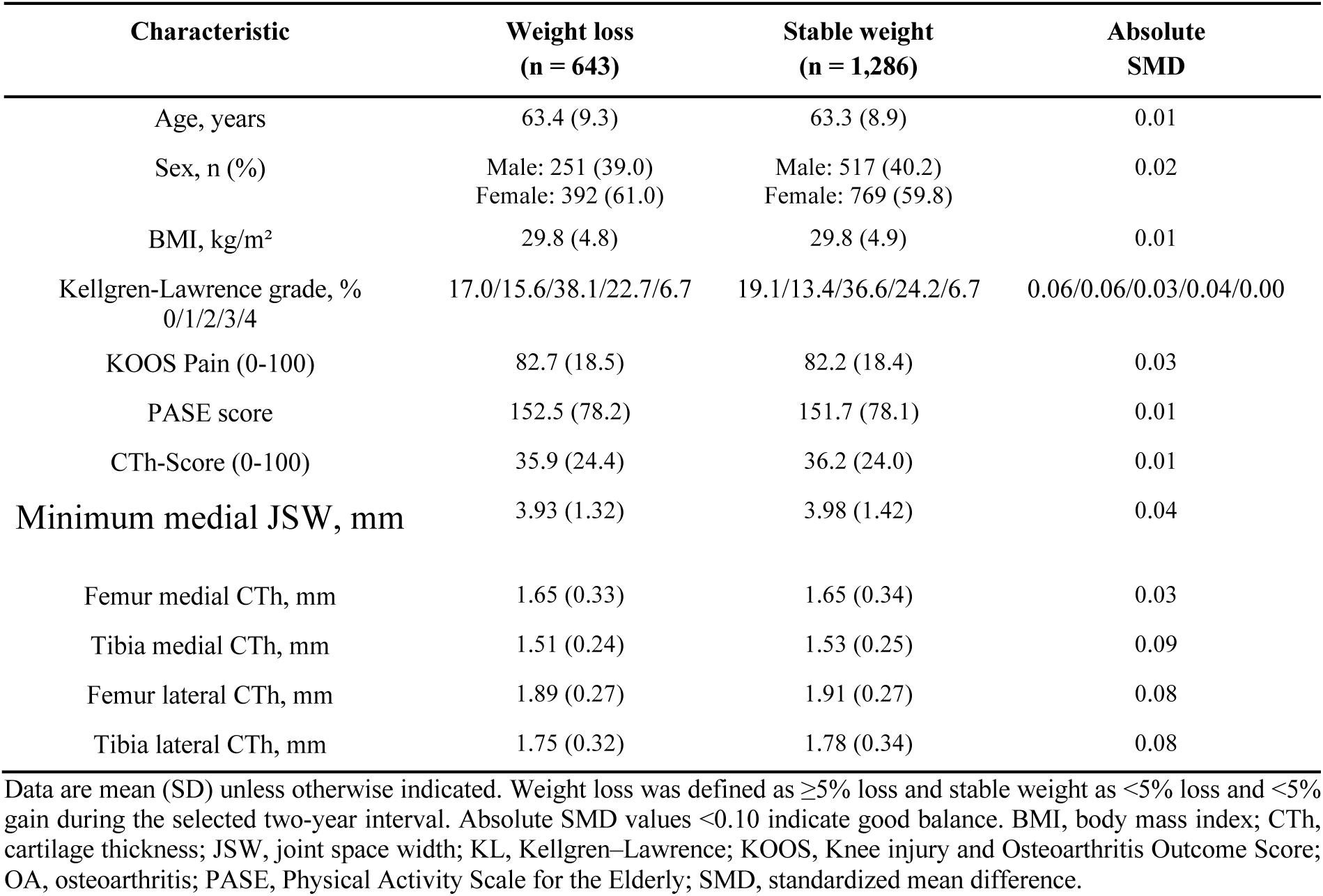
Baseline characteristics by two-year weight-change group.

### Primary Outcome

Participants with weight loss showed less 2-year CTh-Score progression than stable-weight controls (mean change, 2.71 (SD 6.85) vs 4.29 (SD 8.00); Cohen’s d = −0.21) (Table 2). The crude between-group difference was −1.58 points (95% CI −2.28 to −0.89), and the adjusted difference was −1.59 (95% CI −2.29 to −0.89; p<0.001). Adjusted marginal mean changes were 2.70 (95% CI 2.18 to 3.23) and 4.30 (3.86 to 4.73), respectively.

**Table 2.** Two-year outcome changes and between-group estimates.

| Outcome | Two-year change, mean (SD) |  |  | Crude |  | Adjusted |  |
| --- | --- | --- | --- | --- | --- | --- | --- |
|  | Stable weight<br>(n = 1,286) | Weight loss<br>(n = 643) | Cohen's d | Difference (95% CI) | p value | Difference (95% CI) | p value |
| CTh-Score (0-100) | 4.29 (8.00) | 2.71 (6.85) | -0.21 | -1.58 (-2.28 to -0.89) | <0.001 | -1.59 (-2.29 to -0.89) | <0.001 |
| Minimum medial JSW<br>(mm) | -0.16 (0.60) | -0.12 (0.57) | 0.06 | 0.04 (-0.02 to 0.09) | 0.180 | 0.03 (-0.02 to 0.09) | 0.200 |
| Femur medial CTh (mm) | -0.03 (0.12) | -0.03 (0.11) | 0.05 | 0.01 (-0.01 to 0.02) | 0.320 | 0.00 (-0.01 to 0.02) | 0.370 |
| Femur lateral CTh (mm) | -0.02 (0.10) | -0.02 (0.09) | 0.01 | 0.00 (-0.01 to 0.01) | 0.880 | 0.00 (-0.01 to 0.01) | 0.970 |
| Tibia medial CTh (mm) | -0.02 (0.07) | -0.02 (0.07) | 0.01 | 0.00 (-0.01 to 0.01) | 0.760 | 0.00 (-0.01 to 0.01) | 1.000 |
| Tibia lateral CTh (mm) | -0.03 (0.08) | -0.03 (0.08) | -0.03 | 0.00 (-0.01 to 0.01) | 0.580 | 0.00 (-0.01 to 0.00) | 0.470 |
| KOOS Pain (0-100) | 1.45 (14.24) | 2.40 (15.09) | 0.07 | 0.95 (-0.39 to 2.29) | 0.160 | 1.14 (-0.08 to 2.35) | 0.070 |

Changes are mean (SD); differences are weight loss minus stable weight. Adjusted models include the baseline outcome, start age, sex, BMI, radiographic OA severity (KL), visits, KOOS Pain, and PASE, with cluster-robust standard errors by matched set. A negative CTh-Score difference between groups indicates less structural progression; a positive KOOS Pain change indicates less pain. CI, confidence interval; CTh, cartilage thickness; JSW, joint space width; KOOS, Knee injury and Osteoarthritis Outcome Score; OA, osteoarthritis; PASE, Physical Activity Scale for the Elderly; SD, standard deviation.

There was no evidence that the association differed by radiographic OA status (interaction estimate 0.84, 95% CI −0.62 to 2.29; p=0.261).

### Secondary Outcomes

No statistically significant adjusted between-group differences were identified for JSW or mean cartilage thickness in the medial or lateral femoral or tibial regions (Table 2). The adjusted KOOS Pain difference was 1.14 points (95% CI −0.08 to 2.35; p=0.070), with positive values indicating less pain.

## Discussion

This study demonstrates that a significant weight loss of 5% or more over two years is associated with slower structural progression. Applying the novel CTh-Score based on the assessment of cartilage thickness patterns at high resolution we found that the rate of structural progression at two years was lower in participants who lost weight compared with their weight-stable, propensity-matched counterparts. Notably, this protective association was not detectable in this study using traditional metrics such as joint space width (JSW) or mean cartilage thickness in predefined regions, underscoring the enhanced sensitivity of the CTh-Score.

Weight loss was associated with slower structural progression across radiographic OA status, including among participants without radiographic OA. In such individuals, our findings are noteworthy as they provide evidence for the structural benefits of weight loss in individuals at risk of developing radiographic OA, a context where prevention is paramount [26]. This cohort, often asymptomatic, is generally characterized by subtle cartilage damage that are challenging to quantify [2,26,27]. Traditional metrics like JSW lack sensitivity at this stage, as significant narrowing usually manifests later in the disease process (e.g., KL grade 3) [2,27]. The CTh-Score’s ability to detect significant differences in progression stems from its analysis of the spatial distribution of cartilage thickness, captured in the CTh-Maps, at higher resolution than the current standard of averaging thickness values across subregions.

The finding that a 5% or more weight loss is associated with a measurable structural effect over a relatively short 2-year interval suggests that cartilage morphology might be responsive to intervention even before radiographic signs of OA become evident. This could reinforce the potential of weight loss as a primary preventive strategy. An ongoing clinical trial investigating the protective effects of diet-induced weight loss in obese women over 48 months presents an ideal opportunity to prospectively validate these findings, with the CTh-Score serving as a sensitive primary endpoint [13].

In established radiographic OA, our results address a key uncertainty in the literature concerning the timeframe of structural benefits from weight loss. While the symptomatic improvements of weight loss are well-documented [28], prior studies have reported conflicting evidence regarding its concurrent structural impact, with some suggesting that such effects become apparent over longer periods [15,16]. Our findings suggest that structural benefits occur concurrently with weight loss, rather than as a delayed phenomenon. By demonstrating a clear association between weight loss and reduced structural progression within a two-year window, this study provides structural evidence supporting the current clinical guidelines that recommend weight management as a core treatment strategy for patients with knee OA [28].

Achieving and maintaining significant weight loss through diet and exercise remains a major challenge for many individuals. However, the emergence of highly effective pharmacological agents, such as glucagon-like peptide-1 (GLP-1) receptor agonists, offers a promising new therapeutic avenue [29,30]. These agents have demonstrated marked efficacy in inducing substantial weight loss and have shown promise for improving OA symptoms in recent clinical trials [12]. Importantly, emerging evidence suggests that GLP-1 receptor agonists may exert effects on joint tissues that extend beyond their impact on body weight. Several recent studies indicate a potential direct association between GLP-1 and OA, independent of weight loss alone [30]. An open question remains regarding their direct impact on joint structures, potentially extending beyond the secondary effects of weight reduction [30]. Given the demonstrated sensitivity of the CTh-Score in this observational study to detect structural changes linked to weight loss, it represents a valuable tool for future clinical trials. Employing the CTh-Score as an endpoint in trials of GLP-1 agonists could clarify whether these treatments confer a direct chondroprotective effect, thereby contributing to the validation of a novel and potentially potent disease-modifying strategy for OA.

This study has several limitations. As an observational design, it can establish associations but not causality. The specific methods used by participants to achieve weight loss (e.g., diet, exercise, or their combination) were not controlled for, and the reasons why participants lost weight were unknown. This limits the interpretation of the observed associations, as weight loss may reflect heterogeneous underlying mechanisms. In addition, the small effect size observed in CTh-Score change highlights the marked heterogeneity of osteoarthritis progression and indicates that cartilage deterioration cannot be explained by body weight alone, especially in the short term. Pain outcomes should also be interpreted with caution, as baseline KOOS Pain scores were already high, which may have limited the ability to detect further improvement. Finally, while a 2-year follow-up is relevant for clinical trials, longer-term studies are required to determine if these structural benefits are sustained over time.

## Conclusion

In conclusion, this study provides evidence that achieving ≥5% weight loss over 2 years is associated with slower structural progression. The use of the CTh-Score enabled detection of these structural benefits on cartilage where traditional metrics were not significant. These findings strengthen the role of weight loss not only as a cornerstone treatment for managing established OA but also as a critical preventative strategy in at-risk individuals.

## Data Availability

All data produced in the present study are available upon reasonable request to the authors

## Acknowledgements

The authors thank the Osteoarthritis Initiative (OAI) investigators, clinical staff and participants at each of the clinical centres and at the coordinating centre for their important contributions in acquiring the publicly available clinical and imaging data. The OAI is a public–private partnership comprising five contracts (N01-AR-2-2258; N01-AR-2-2259; N01-AR2-2260; N01-AR-2-2261; N01-AR-2-2262) funded by the NIH and conducted by the OAI Study Investigators. Private funding partners of the OAI include Merck Research Laboratories, Novartis Pharmaceuticals Corporation, GlaxoSmithKline, and Pfizer. Private sector funding for the OAI is managed by the Foundation for the NIH.

## Contributions

PM, JF, and PO conceived and designed the study. JF and PO supervised the study and secured funding. PM collected and assembled the data. PM, JF, and PO analyzed and interpreted the data. PM and PO drafted the article. FB contributed to interpretation and reviewed the manuscript. All authors critically revised the article for significant intellectual content and approved the final version.

## Role of the funding source

This work was funded by the Swiss National Science Foundation, Switzerland (SNSF Grant#CRSII5_177155 & CRSII--222725). The funding sources had no role in the study design, collection, analysis, and interpretation of data; in the writing of the manuscript, and in the decision to submit the manuscript for publication.

## Conflict of interest

PM, JF, and PO have no conflicts of interest. FB reports consulting fees from Grunenthal, GSK, Eli Lilly, Novartis, Pfizer, Servier, and 4P Pharma; honoraria from Viatris, Pfizer, and Zoetis; travel support from Nordic Pharma; patents with 4Moving Biotech; advisory board/DSMB roles with AstraZeneca, Sun Pharma, and Nordic Bioscience; and stock/options in 4Moving Biotech and 4P Pharma.

## Notes

### Competing Interest Statement

The authors have declared no competing interest.

### Summary of Updates

Redesign of the study following submission to OAC

